# Does autism protect against COVID quarantine effects?

**DOI:** 10.1101/2020.10.13.20212118

**Authors:** Marco Guidotti, Adrien Gateau, Joelle Malvy, Frédérique Bonnet-Brilhault

## Abstract

**Introduction:** COVID-19 outbreak has imposed an eight-week confinement in France. During this period, children and their families were exposed to a full-time home life. The aim of this study was to assess the emotional experience and tolerance of children with autism spectrum disorder (ASD) in this particular context.

**Method:** A clinical survey was proposed to parents and rated by professionals once a week during the quarantine period in France. 95 autistic children followed by the child and adolescent psychiatry department of Tours university hospital were assessed from the 18th of March to the 8th of May. The following clinical points were investigated: child anxiety, family anxiety, behavior problems, impact on sleep, impact on appetite, impact on school work, family tension, confinement intolerance, difficulties to follow a schedule, isolation behavior.

**Results:** Despite minor changes in family anxiety and school work, no difference was highlighted between clinical scores collected at the beginning and at the end of this period. ASD children with or without intellectual disability had non-significant clinical changes during quarantine. This evolution was also independent of the accommodation type (house or apartment) and the parental status (relationship, separated or isolated).

**Conclusion:** The sameness dimension in autism and parents’ adaptation may be involved in this clinical stability during COVID confinement. Moreover, specialized tools and support provided by professionals could have participated to these outcomes and must be regularly promoted in order to help families in this still difficult period.

## Introduction

COVID-19 outbreak began in China at the end of 2019 and quickly spread to Europe. In France, the confinement of the population began on the 17^th^ of March and schools closed the day before. This new situation, which lasted officially until the 11^th^ of May, exposed children and parents to a full-time home life. We know that this lifestyle can be difficult for families, especially for children suffering from mental diseases [1]. The psychological impact of quarantine is heterogeneous and can be long lasting, including anxiety, depression symptoms and feelings of anger and fear. To limit these consequences, it is recommended to reduce boredom and improve communication [2]. In order to help parents at home, some advice was published to manage children suffering from autism spectrum disorder (ASD) [3]. Furthermore, health professionals regularly contacted families to get news and to provide some personal advice. Considering these clinical resources, we investigated how well the COVID-19 quarantine was tolerated by autistic children who benefit from regular rehab programs in the child and adolescent psychiatry department of Tours university hospital.

## Material and methods

A telephone survey was proposed to families, once a week, during French lockdown. The following clinical points were investigated by health professionals (doctors, nurses and psychologists): child anxiety, family anxiety, behavior problems, impact on sleep, impact on appetite, impact on school work, family tension, confinement intolerance, difficulties to follow a schedule, isolation behavior. Each of these ten clinical items was rated on a 5-point scale in reference to the basal clinical state of the child: 1/not at all, 2/a little, 3/moderately, 4/a lot or 5/enormously. The mean of the completed clinical items, labelled emotional score, was computed. Two time points were considered: the 3 first weeks (T1) and the 3 last weeks of the recruitment period (T2). Furthermore, the accommodation type (apartment or house) and the parental status were also collected.

Additionally, parents were regularly contacted by professionals and many supporting materials were sent to families (sensorimotor activities, timetables, simplified COVID explanations) during this period.

SphinxOnline v 4.16 was used for data collection. Student t-test and linear mixed-effect models with repeated measures (patients as random effect; time and ID, accommodation type or parental status as fixed effects) were carried out to study the effect of the quarantine between T1 and T2 time points.

Approval of Tours hospital ethical committee has been granted to conduct this project.

## Results

95 children with ASD, aged from 2 to 16 years old, were recruited from the 18^th^ of March to the 8^th^ of May in the child and adolescent psychiatry department of Tours university hospital (Table 1).

**Table 1:** Characteristics of the sample

|  |  |  |
| --- | --- | --- |
| Sample (children) |  | 95 |
| Age (years) : Mean +/- SD |  | 7,1 +/- 3,3 |
| Gender | Male | 78 (82.1%) |
|  | Female | 17 (17.9%) |
| Comorbidities | Intellectual Disability | 32 (33.7%) |
|  | Language disorders | 12 (12.6%) |
|  | Motor disorders | 7 (7.4%) |
|  | ADHD | 1 (1.0%) |
| Living place | House | 37 (39%) |
|  | Apartment | 58 (61%) |
| Parental status | Relationship | 63 (66.3%) |
|  | Separated | 19 (20.0%) |
|  | Isolated | 13 (13.7%) |

Eight clinical items (child anxiety, behavior problems, impact on sleep, impact on appetite, family tension, confinement intolerance, difficulties to follow a schedule, isolation behavior) had an average score of less than 2 and two items (family anxiety and impact on school work) were greater than 2 at both T1 and T2 time points (Fig.1).

**Fig 1.**
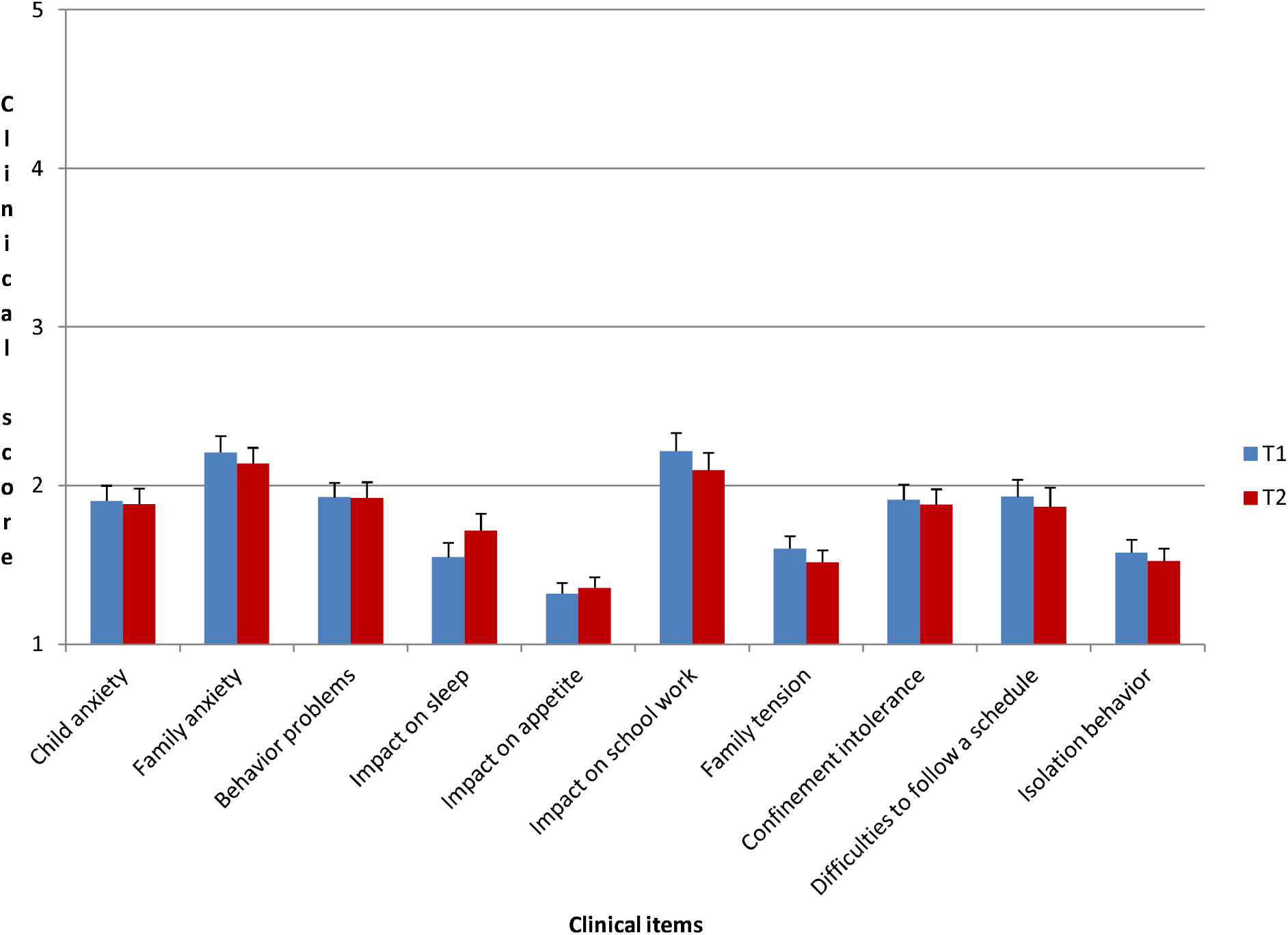
Clinical items (mean ± sem) at T1 and T2 time points (N=95)

The emotional score (N=95) was 1.82 (SD: 0.59) at T1 and 1.78 (SD: 0.59) at T2, no statistical difference was observed (t(188)=0.44, p=0.66)). Moreover there was no DI effect (DI effect: p=0.54, interaction DI * time: p=0.83), no accommodation type effect (accommodation type effect: p=0.12, interaction accommodation type * time: p=0.61), and no parental status effect (parental status effect: p=0.88, interaction parental status * time: p=0.93).

## Discussion

No difference was highlighted between clinical scores collected at the beginning and at the end of French COVID confinement in our sample. Despite minor changes in family anxiety and impact on school work, the emotional score was unchanged. This evolution was not found by Colizzi et al. (2020), who highlight more frequent and intense behavior problems during confinement [4]. However, unlike Colizzi’s study, our investigation was a prospective survey and included children followed by professionals. The latter regularly contacted families and offered adapted and personalized tools during quarantine. Indeed, we argue that the materials sent and regular support provided by professionals prevented an increase of anxiety and behavioral disturbances. Our outcomes may also be linked to autism’s sameness dimension [5], associated to the tendency to seek immutability and to be afraid of changes [6]. Furthermore, the decrease of social interactions and sensory exterior inputs during the confinement could have helped to maintain environmental stability.

Another result of our survey was the minor changes in family anxiety and school work. During quarantine, parents had to ensure their children’s schooling in addition to their work. This double task was complicated, especially since the education of autistic children presents several particularities and needs specific materials [7]. Moreover, Lei et al. (2020) described an increased prevalence of anxiety and depression in adults affected by quarantine [8], which could have participated to our parents’ distress.

Furthermore, we found that the accommodation type and parental status did not influence the emotional experience and tolerance of children to quarantine. These independent factors can highlight the significance of routine and environmental stability. On the other hand, autistic children were allowed to go out longer (> 1 hour per day) than other children from the 2^nd^ of April (from the mid-time of quarantine); this political decision could have impacted the effect of these variables.

Finally, our survey has some limits. Firstly, clinical points were evaluated by parents and rated by professionals who know the children: this method does not ensure the uniformity of measurements. Secondly, our study was based on children followed by a single center with specific procedures which limits generalization to other situations.

## Conclusion

This prospective study underlines the unchanged emotional experience and behavior in ASD children during quarantine in France. ASD particularities and parents’ adjustments to the situation probably contributed to this clinical stability; moreover, professionals’ support and specialized tools likely participated to these results as well and must be promoted in order to help families in this still difficult period. Finally, going back to school and the resumption of activities with COVID restrictions will be fragile moments in ASD children’s lives, and would deserve a specific assessment.

## Data Availability

Due to their confidentiality, the clinical data might be obtained upon request by contacting the corresponding author.

## References

1. Asbury K, Fox L, Deniz E, Code A, Toseeb U. How is COVID-19 Affecting the Mental Health of Children with Special Educational Needs and Disabilities and Their Families? J Autism Dev Disord. Published online July 31, 2020:1–9. doi:10.1007/s10803-020-04577-2

2. Brooks SK, Webster RK, Smith LE, et al. The psychological impact of quarantine and how to reduce it: rapid review of the evidence. The Lancet. 2020;395(10227):912–920. doi:10.1016/S0140-6736(20)30460-8

3. Cluver L, Lachman JM, Sherr L, et al. Parenting in a time of COVID-19. Lancet Lond Engl. 2020;395(10231):e64. doi:10.1016/S0140-6736(20)30736-4

4. Colizzi M, Sironi E, Antonini F, Ciceri ML, Bovo C, Zoccante L. Psychosocial and Behavioral Impact of COVID-19 in Autism Spectrum Disorder: An Online Parent Survey. Brain Sci. 2020;10(6). doi:10.3390/brainsci10060341

5. Szatmari P, Georgiades S, Bryson S, et al. Investigating the structure of the restricted, repetitive behaviours and interests domain of autism. J Child Psychol Psychiatry. 2006;47(6):582–590. doi:10.1111/j.1469-7610.2005.01537.x

6. Diagnostic and Statistical Manual of Mental Disorders: DSM-5TM, 5th Ed. American Psychiatric Publishing, Inc.; 2013:xliv, 947. doi:10.1176/appi.books.9780890425596

7. Marsh A, Spagnol V, Grove R, Eapen V. Transition to school for children with autism spectrum disorder: A systematic review. World J Psychiatry. 2017;7(3):184–196. doi:10.5498/wjp.v7.i3.184

8. Lei L, Huang X, Zhang S, Yang J, Yang L, Xu M. Comparison of Prevalence and Associated Factors of Anxiety and Depression Among People Affected by versus People Unaffected by Quarantine During the COVID-19 Epidemic in Southwestern China. Med Sci Monit Int Med J Exp Clin Res. 2020;26:e924609. doi:10.12659/MSM.924609

